# Genetics of Motor Neuron Disease in New Zealand

**DOI:** 10.1101/2024.09.24.24314000

**Authors:** Miran Mrkela, Miriam Rodrigues, Serey Naidoo, Jules B.L. Devaux, Siobhan E. Kirk, Chitra Vinnakota, Christina M. Buchanan, Dympna Mulroy, Harry Fraser, Jessie C. Jacobsen, Hannah Wyatt, Kylie Drake, Elsa Parker, Howard Potter, Lyndal Henden, Emily P. McCann, Kelly L. Williams, Anjali K. Henders, Richard H. Roxburgh, Emma L. Scotter

**Affiliations:** School of Biological Sciences, The University of Auckland, Auckland, New Zealand; Centre for Brain Research, The University of Auckland, Auckland, New Zealand; Neurology Department, Te Toka Tumai, Te Whatu Ora, Auckland, New Zealand; Canterbury Health Laboratories, Te Whatu Ora, Waitaha Canterbury, Christchurch, New Zealand; Macquarie University Centre for Motor Neuron Disease Research, Faculty of Medicine, Health and Human Sciences, Macquarie University, Sydney, NSW, Australia; The University of Queensland, Queensland Brain Institute, Brisbane, QLD, Australia; Neurogenetics Research Clinic, School of Medicine, Faculty of Medical and Health Sciences, University of Auckland, Auckland, New Zealand

**Keywords:** Amyotrophic lateral sclerosis, motor neuron disease, *C9orf72*, *SOD1*, genetic screening, New Zealand, pre-symptomatic, genetics

## Abstract

Motor neuron disease (MND) is a group of neurodegenerative diseases characterised by adult-onset progressive degeneration of motor neurons, of which amyotrophic lateral sclerosis (ALS) is the most common. MND is clinically heterogenous with complex aetiology, being caused by or associated with over 40 different genes, along with multiple environmental risk factors. Although New Zealand has one of the highest incidence and mortality rates of MND globally, the reasons for this are yet to be determined.

In light of this lack of systematic genetic MND research in New Zealand, we sought to identify the frequencies of genetic variants in known MND-linked genes in people in New Zealand with MND. A total of 184 participants were enrolled; 149 with a clinical diagnosis of MND and 35 who were clinically unaffected but at risk of familial MND. Of the 149 affected, 128 had sporadic MND, and 21 had familial MND. Participants’ DNA was screened for genetic variation in 46 MND-associated genes using a combination of Sanger sequencing, Illumina single nucleotide polymorphism (SNP) microarray, repeat-primed PCR for *C9orf72*, and an Invitae gene panel.

Clinical phenotypes of participants with MND recapitulated the trends seen in European reference populations with both males, and individuals with spinal onset, having earlier disease onset. Thirty three of the 184 participants (17.9%) carried known pathogenic variants; the majority had *C9orf72* hexanucleotide repeat expansion (24/34), and the remainder had previously reported pathogenic genetic variants in *SOD1* (9/34; p.(Ile114Thr) and p.(Glu101Gly)). All NZ *SOD1* p.(Ile114Thr) cases (n=4) were distantly related (identical-by- descent) to one another and a cluster of >30 MND cases from Australia with the same variant. Variants of interest were identified in 14 participants, of which splicing variants *DCTN1*:c.279+1G>C and *ATP13A2* p.(Lys804=) are subject to ongoing study. Of the pathogenic variants, 48.4% were identified in pre-symptomatic unaffected individuals with a family history, highlighting the importance of offering cascade testing and symptom surveillance for families, particularly as gene-specific treatments are becoming available.

## 1.1 Introduction

Motor neuron disease (MND) is a group of neurodegenerative diseases characterised by adult-onset progressive degeneration of motor neurons (Hardiman et al., 2017). Amyotrophic lateral sclerosis (ALS), characterised by the selective degeneration of both upper and lower motor neurons (Shatunov and Al-Chalabi, 2021), is the most common form of MND, comprising 80-90% of MND diagnoses (Roman, 1996; Rowland and Shneider, 2001). Globally, New Zealand has among the highest mortality and incidence rates (Cao et al., 2018; Logroscino et al., 2018; Park et al., 2022), yet no large-scale genetic studies have been undertaken in New Zealand to determine whether there is a genetic basis for this high rate of MND. New Zealand also lacks standardisation in MND-specific clinical genetic testing.

ALS and MND are commonly binary-classified into familial (fMND) and sporadic (sMND) subtypes based on observations of familial clustering (Al-Chalabi and Hardiman, 2013; Renton et al., 2014). However, there is increasing evidence to suggest that these subtype distinctions are largely artificially constructed and that it is a combination of genetic predisposition and environmental factors that lead to disease manifestation in both subtypes (Al-Chalabi et al., 2014). The aetiology of MND is complex; to date, more than 40 genes are directly causative of, or are associated with, the ALS subtype (Abel et al., 2012; Landrum et al., 2018; Shatunov and Al-Chalabi, 2021). Environmental factors including cigarette smoking, agricultural work, and electrocution injury (Chen et al., 2021, 2019; Vasta et al., 2022) have also been identified as risk factors. Despite the identified causal genetic associations, only 5-10% of people with ALS have a Mendelian family history of disease, with the majority of cases appearing in families where seemingly only a single individual is affected (sporadic) (Al-Chalabi et al., 2014; Al-Chalabi and Lewis, 2011).

The majority of New Zealanders (∼70.2%) are of European ethnicity (Stats NZ–Tatauranga Aotearoa, 2018) so it is likely that genetic population trends reported in European populations are reflected at least partially in New Zealand. In European populations, the most commonly implicated genes in people with MND are *C9orf72* (∼40% fMND | 7-10% sMND), *SOD1* (∼13-25% fMND | 1-2% sMND), *TARDBP* (2-4% fMND | <1% sMND), and *FUS* (2-4% fMND | <1% sMND) (Brown et al., 2021; Majounie et al., 2012; McCann et al., 2017; Millecamps et al., 2010; Renton et al., 2014; Zou et al., 2017). Thus, testing New Zealanders for only these four genes could theoretically identify approximately 60-73% of genetic causes of fMND and 8-12% of sMND in a cost effective and standardised manner. However, as New Zealand also has individuals who identify as Māori, Asian, or Pacific peoples (16.5%, 15.1%, and 8.1% respectively) there may be population-dependent genetic variation contributing to high MND rates in New Zealand (Stats NZ–Tatauranga Aotearoa, 2018). Given the lack of genetic research of MND in New Zealand, and the ethnically diverse population, we sought to identify the primary genetic causes of MND in New Zealanders by using a combination of Sanger sequencing, Illumina SNP microarrays, repeat-primed PCR, and a commercial Invitae gene panel to test for genetic variants that are causative of, or associated with, MND, in a total of 46 genes. This study identified pathogenic variants and variants of uncertain significance in both clinically affected and at-risk individuals.

## 1.2 Methods

### 1.2.1 Study participants and consent

The study was approved by the Health and Disability Ethics Committee of New Zealand (HDEC approval 19/CEN/7). The New Zealand cohort consisted of participants recruited from throughout New Zealand (Supplementary Fig. 1) through general practitioners, neurologists, self-referral, or the New Zealand Motor Neurone Disease Registry (https://mnd.org.nz/research/mnd-registry/). All participants provided informed written consent after telephone consultation with a genetic counsellor. Inclusion criteria were: having a verified diagnosis of MND by a neurologist (‘affected’); or being a first- to third-degree relative of a person with familial MND (‘unaffected family’). Familial MND (fMND) was defined as having a verified diagnosis of MND and having one or more first- to second- degree relatives also with a verified diagnosis of MND (Byrne et al., 2011). Sporadic MND (sMND) was defined by the absence of such family history. In general, New Zealand clinicians do not differentiate between MND subtypes when making a formal diagnosis so affected participants in our cohort were not analysed by MND subtype. All family and participant IDs used within this study are non-identifying and known only to select research members.

### 1.2.2 Disease phenotype analysis

The following disease phenotype parameters: MND inheritance (fMND or sMND), sex, and site of onset (bulbar or spinal), were examined in the New Zealand cohort and a larger European MND cohort from the Project MinE Consortium. The same clinical parameters for the New Zealand cohort was taken from the New Zealand Motor Neurone Disease Registry. However, not all participants had all data available.

For the European MND cohort, clinical information was available for 2792 MND cases provided by the Project MinE ALS Sequencing Consortium. All ALS cases were diagnosed with definite or probable ALS according to the revised El Escorial criteria (Brooks et al., 2000). All cases were then classified as either ‘familial’ or ‘sporadic’, with ‘familial’ cases being those that satisfied the requirements for probable or definite fMND (Byrne et al., 2011).

Clinical parameters and age of disease onset were compared statistically within each cohort and between cohorts using pair-wise Wilcoxon rank sum tests with Bonferroni multiple comparisons adjustment (α: 4.2×10^-3^). All statistical analysis was conducted in R (v.4.0.5) using the packages: tidyverse, ggpubr, Cairo, and data.tables (Dowle and Srinivasan, 2021; Kassambara, 2020; Urbanek and Horner, 2022; Wickham et al., 2019).

### 1.2.3 Genetic analysis

Genomic DNA was extracted by Canterbury Health Laboratories from peripheral blood (EDTA) or saliva (Oragene 500) using a Kurabo QuickGene DB-L salt extraction-based kit or Oragene PrepIT method, respectively. DNA aliquots underwent commercial diagnostic testing by Invitae and/or research testing by the University of Queensland. The genetic analysis workflow utilised an established sequencing pipeline at the University of Queensland. Affected participants could choose whether they received their genetic results (confirmed in a certified clinical laboratory); no predictive results (of unaffected participants) were returned.

### 1.2.3.1 Sanger sequencing and SNP microarray

Sanger sequencing and SNP microarray were conducted at the University of Queensland. Sanger sequencing was used for the first 50 participants to screen for 12 common pathogenic variants in *SOD1 (OMIM: 147450), FUS (OMIM: 137070), TARDBP (OMIM: 605078),* and for risk alleles in *C9orf72 (OMIM: 614260;* rs3849942) and *UNC13A (OMIM: 609894)* rs12608932 (Supplementary Table 1). Sanger sequencing for these variants was later replaced by microarray genotyping (n=135 participants). *SOD1* rs121912439, NM_000454.5:c.302A>G p.(Glu101Gly) continued to be assessed by Sanger sequencing as it was not represented on the microarray.

SNP microarray genotyping used the Illumina Infinium High Throughput Screening (HTS) Assay (Global screening array, GSA) with GSAv3.0 BeadChips. Standard Illumina Infinium HTS protocols were used with loading and staining performed by a Tecan Freedom Evo robot. BeadChips were scanned using the Illumina iScan within 24 h of staining. Illumina Genome Studio v2.0 software was used to analyse iScan data, using Illumina bead-pool manifest file GSAMD-24v3-0-EA_20034606_B1.bpm. Call rates were generated for each sample with >0.99 considered ideal and <0.95 considered failed.

The 654,027 SNP loci represented on the BeadChip were intersected with 853 previously reported MND-associated variants (McCann et al., 2020), indicating that 51 MND SNPs (causative or associated with) across 18 genes were represented on the BeadChip (Supplementary Table 2). DNA samples carrying variants deemed ‘likely pathogenic’ or ‘pathogenic’ according to American College of Medical Genetics (ACMG) classifications were re-tested by Canterbury Health Laboratories using accredited PCR and bidirectional fluorescent DNA sequencing, before return of validated genetic results to study participants by the consenting genetic counsellor.

Illumina GSA data was also used to evaluate genetic ancestry to identify the most relevant Genome Aggregation Database (gnomAD) reference population for SNP minor allele frequencies (MAFs). Briefly, GSA data was merged with HapMap 3 data (The International HapMap Consortium, 2003) using a combination of PLINK and R (v.4.0.5) following a pipeline set out by (Meyer, 2020). SNPs with a MAF of <0.01 or a variant inflation factor >2 were filtered out. Principal component analysis (PCA) was conducted using KING (Manichaikul et al., 2010) and the result plotted using R (v.4.0.5).

#### 1.2.3.2 Commercial Invitae diagnostic high throughput sequencing panel

DNA from individuals: i) with fMND or young-onset (<35 y) sMND, in whom we had not identified a pathogenic MND variant by Sanger, SNP array, or *C9orf72* repeat expansion analysis; or ii) with an affected first-to-seventh-degree relative by identity-by-descent (IBD) analysis, was sequenced using a commercial NGS panel of 42 ALS, FTD or Alzheimer disease-related genes by Invitae (n=29) (Test code 03502, Supplementary Table 3). The protocol for sequencing has been described before (Lincoln et al., 2015) but briefly, targeted genes were captured using SureSelect (Agilent, Santa Clara, CA) or Integrated DNA Technologies (Coral, IL) probes. NGS was performed either on the Illumina (San Diego, CA) MiSeq or HiSeq 2500 to at least 450x average coverage of 150-bp paired-end reads, with a minimum of 50x required at every targeted position (Lincoln et al., 2015).

Reads were aligned to the GRCh37 reference genome and genetic variants were annotated focusing on exonic regions, 20 bp of flanking intronic sequence, and regions demonstrated to be either causative or associated with ALS, FTD, or Alzheimer’s disease at the time of assay design. Variant interpretation was conducted by Invitae using the Sherloc variant interpretation framework (Nykamp et al., 2017) and a report generated for each individual summarising variants of uncertain significance, likely pathogenic, and pathogenic variants. Benign or likely benign variants were not reported.

#### 1.2.3.3 *C9orf72* repeat expansion analysis

Repeat-primed PCR was conducted by Canterbury Health Laboratories to test for the *C9orf72* GGGGCC hexanucleotide repeat expansion (NM_001256054.1(C9orf72):c.- 45+163GGGGCC[>60]). Briefly, two PCR reactions were undertaken: the first encompassing the repeat to calculate the number of normal/intermediate repeats, and the second to detect large expansions by variably priming the repeat region (see Supplementary Table 4). PCR products were analysed by fragment analysis using fluorescently labelled products separated by capillary electrophoresis on the ThermoFisher ABI3500XL. Repeat classifications were set as follows: Normal: <20 repeats, intermediate: 20-60 repeats, and pathogenic: ≥60 repeats, except for three participants who had had historical testing when ≥30 repeats was defined to be pathogenic and for whom DNA was not available to re-test (Iacoangeli et al., 2019).

### 1.2.4 *In silico* variant pathogenicity prediction

The pathogenicity of missense and splice site variants of uncertain significance identified by GSA or Invitae panel was examined using *in silico* pathogenicity prediction algorithms. To determine optimal algorithms for MND variant predictions, ClinVar (https://www.ncbi.nlm.nih.gov/clinvar/) was mined for variants associated with “Amyotrophic Lateral Sclerosis” and with at least one star of evidence (n=1,244 variants). Benign variants were defined as either “likely benign” or “benign” variants and similarly, pathogenic variants were defined as either “likely pathogenic” or “pathogenic” and were labelled as the variant ‘ground truth’. These were converted to .vcf format and parsed through predictive algorithms on dbNSFP (http://database.liulab.science/dbNSFP) (Dong et al., 2015; Liu et al., 2020). A total of 273 variants remained which had entries for variant predictions. Classification values were set according to (Supplementary Table 5). Variant predictions were plotted against ‘ground-truth’ variants (Supplementary Fig. 2A) and Cohen’s Kappa computed to evaluate predictor accuracy and specificity (Supplementary Fig. 2B and Fig. 2C).

Based on those results; for missense variants, the meta-predictors Combined Annotation Dependent Depletion (CADD) (Rentzsch et al., 2019), Rare Exome Variant Ensemble Learner (REVEL) (Ioannidis et al., 2016), BayesDel (Tian et al., 2019), and MetaRNN (Li et al., 2022) were used to predict deleteriousness. Variants that did not meet a conservative CADD threshold of 15 were not included for further analysis. All variants were also parsed through CI-SpliceAI (Strauch et al., 2022) and SPiCEv2.1/SPiP (Leman et al., 2022, 2018) for predicting altered splicing.

### 1.2.5 Identity-by-descent analysis of participants with *SOD1*

Pairwise identity-by-descent (IBD) analysis was performed to determine whether New Zealand participants with a *SOD1* NM_000454.5:c.341T>C, p.(Ile114Thr) variant were genetically related to Australian ALS cases with the same variant (Henden et al., 2020) - therefore sharing a disease haplotype with a common founder. Forty two *SOD1* p.(Ile114Thr) ALS participants were selected for IBD analysis (New Zealand, n=4; Australia, n=38). Whole-genome sequencing (WGS) data was available for 4 samples, while SNP microarray data was available for 38 samples genotyped on either the Infinium CoreExome array or the GSA array. The WGS and SNP microarray data were merged to extract biallelic SNPs common to all platforms, resulting in 141,212 SNPs for analysis.

Pairwise IBD analysis was performed between all 42 individuals using XIBD (Henden et al., 2016). Briefly, XIBD uses a hidden Markov model and unphased genotype data to identify genomic regions with either one or two alleles shared IBD between pairs of individuals (Henden et al., 2016). A European ancestry reference population was used (samples populations CEU and TSI from HapMap-3, https://www.broadinstitute.org/medical-and-population-genetics/hapmap-3) to calculate linkage disequilibrium and MAF statistics. SNPs in high LD (PLINK squared correlation >0.8) or with MAF <0.01 were removed, resulting in 117,835 SNPs for IBD analysis. IBD segments of 2 cM or larger were retained for further exploration. For each pair of samples, the degree of relationship was estimated using the lengths of inferred IBD segments as in KING (Manichaikul et al., 2010). Networks of related samples were drawn in R (version 3.5.1) using the packages igraph and ggnetwork, with nodes placed according to the Fruchterman-Reingold force-directed layout algorithm (Fruchterman and Reingold, 1991), which aims to position nodes such that all edges are of similar lengths with as few edges overlapping as possible. There is no correlation between the length of the edges and the degree of relatedness.

## 1.3 Results

### 1.3.1 Cohort demographics and disease phenotypes

The cohort for analysis of the genetics of MND in New Zealand comprised 184 participants (50.2% female) from 162 different families (Table 1). Of these 184, 149 had diagnosed MND; 128 (86%) of these had sMND and 43% of these sMND participants were female. Twenty one (14%) from 12 families had fMND and 61.9% of these were female. The remaining 35 were unaffected family members of affected participants (68.6% female). Here, ‘unaffected’ means not diagnosed with MND; this group therefore includes a mixture of gene-negative and pre-disposed gene-positive individuals. Most affected participants (69.6%) presented with spinal onset, 28.8% presented with bulbar onset, and 1.6% with respiratory (chest/abdominal) onset (Table 1). The average age of onset was 50.7 y for familial (SD, 14.3; IQR, 14) and 59.2 y for sporadic (SD, 11.9; IQR, 15).

**Table 1.** Cohort demographics and phenotype characteristics.

| Table 1. Cohort demographics and phenotype characteristics |  |  |  |  |
| --- | --- | --- | --- | --- |
|  | Unaffected family | Sporadic | Familial | Total |
| Total participants: | 35 | 128 | 21 | 184 |
| Sex at birth |  |  |  |  |
| Female (%) | 24 (68.6) | 56 (43.7) | 13 (61.9) | 93 (50.2) |
| Male (%) | 11 (31.4) | 73 (56.3) | 8 (38.1) | 92 (49.7) |
| Age of onset (years) |  |  |  |  |
| Mean [SD] | - | 59.6 [11] | 50.7 [14.3] | 58.5 [12.3] |
| Median | - | 60 | 55 | 59 |
| IQR | - | 15 | 14 | 15 |
| Participants (n) | - | 122 | 13 | 135 |
| Disease duration (months) |  |  |  |  |
| Mean [SD] | - | 3.6 [2] | 10.6 [8.6] | 4.4 [4.1] |
| Median | - | 3.1 | 9.2 | 3.1 |
| IQR | - | 2 | 10.9 | 2.4 |
| Participants (n) | - | 44 | 6 | 50 |
| Site of onset |  |  |  |  |
| Bulbar | - | 31% | 9.1% | 28.8% |
| Spinal | - | 67.3% | 90.9% | 69.6% |
| Respiratory | - | 1.8% | 0.0% | 1.6% |
| Participants (n) | - | 113 | 11 | 124 |

Disease phenotypes of the cohort (n=124 for whom data were available) were compared with a Project MinE European reference cohort (n=2792) (Fig. 1). In the New Zealand cohort, no differences were found in the age of onset between sexes, familial status, or anatomical site of onset (Fig. 1A). In contrast, in the larger Project MinE cohort, differences in the mean age at disease onset were observed between; sexes (Fig. 1B_2_, male 60.6 y, female 63.1 y, Wilcoxon rank sum, p.adj= 4.6×10^-10^) and sites of onset (Fig. 1B_3_, spinal 60.9 y, bulbar 63.6 y, Wilcoxon rank sum, p.adj=7.6×10^-12^), but not between sMND and fMND (Fig. 1B_1_, Wilcoxon rank sum, p.adj=0.112). Additionally, no differences were found between the European reference cohort and the NZ cohort for any of the metrics tested.

**Figure 1.**
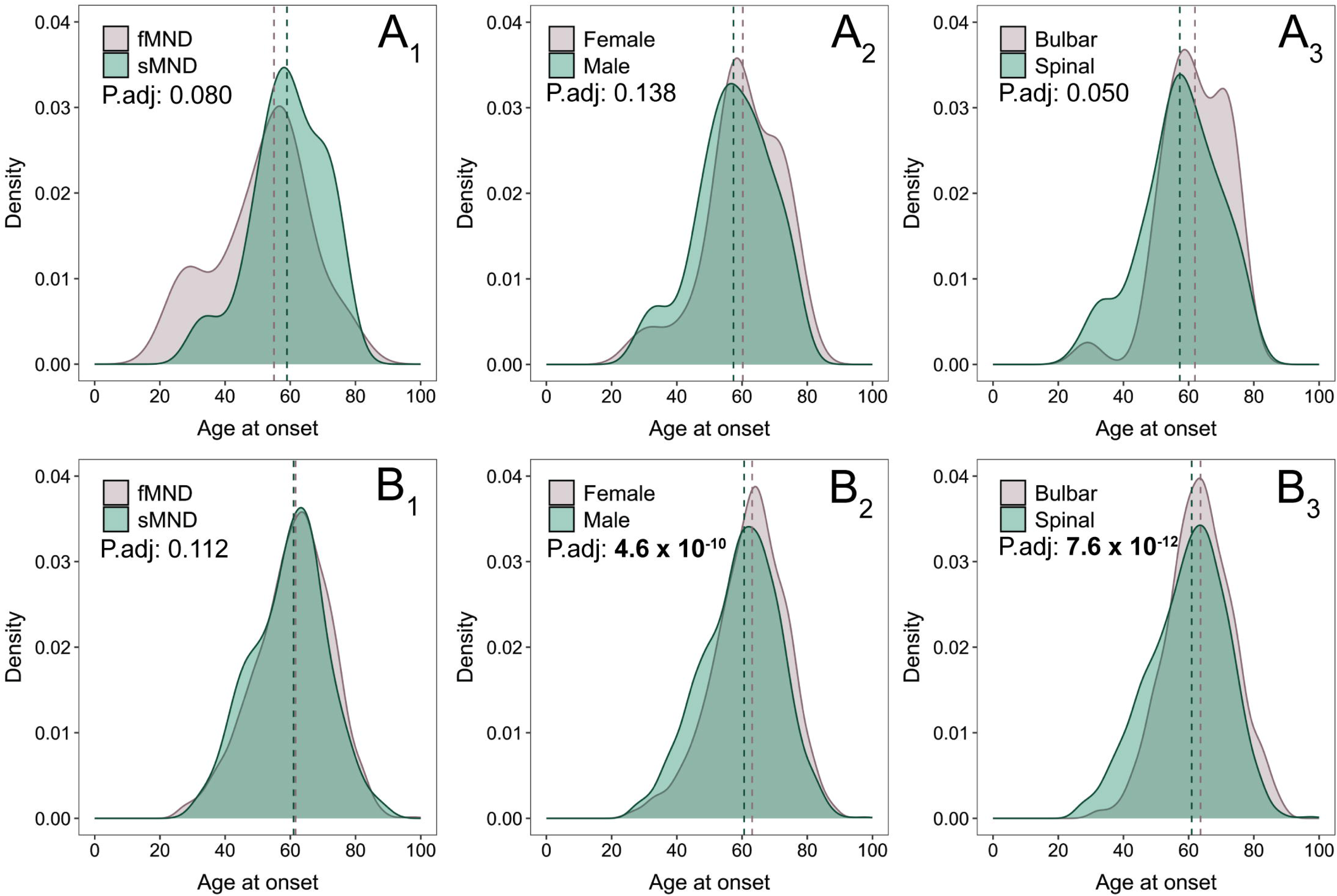
Disease phenotypes of the NZ MND genetics study cohort and European MND reference population. Disease phenotypes of the New Zealand MND genetic study cohort (n=124) were compared with a larger Project MinE European reference MND cohort (n=2792). In the New Zealand cohort (A), no differences were found in age at onset according to family history (A_1_), sex (A_2_), or site of onset (A_3_). In contrast, the larger Project MinE cohort (B) did not identify a difference in age at disease onset between family history (B_1_) but did between sex (males 60.6 y, females 63.1 y, P.adj = 4.6X10^-10^) (B_2_), and site of onset (spinal 60.9 y, bulbar 63.6 y, P.adj = 7.6X10^-12^) (B_3_). Dotted lines denote the respective mean age at disease onset within each panel. All statistical tests were conducted using Wilcoxon rank sum with Bonferroni multiple comparisons.

**Figure 2.**
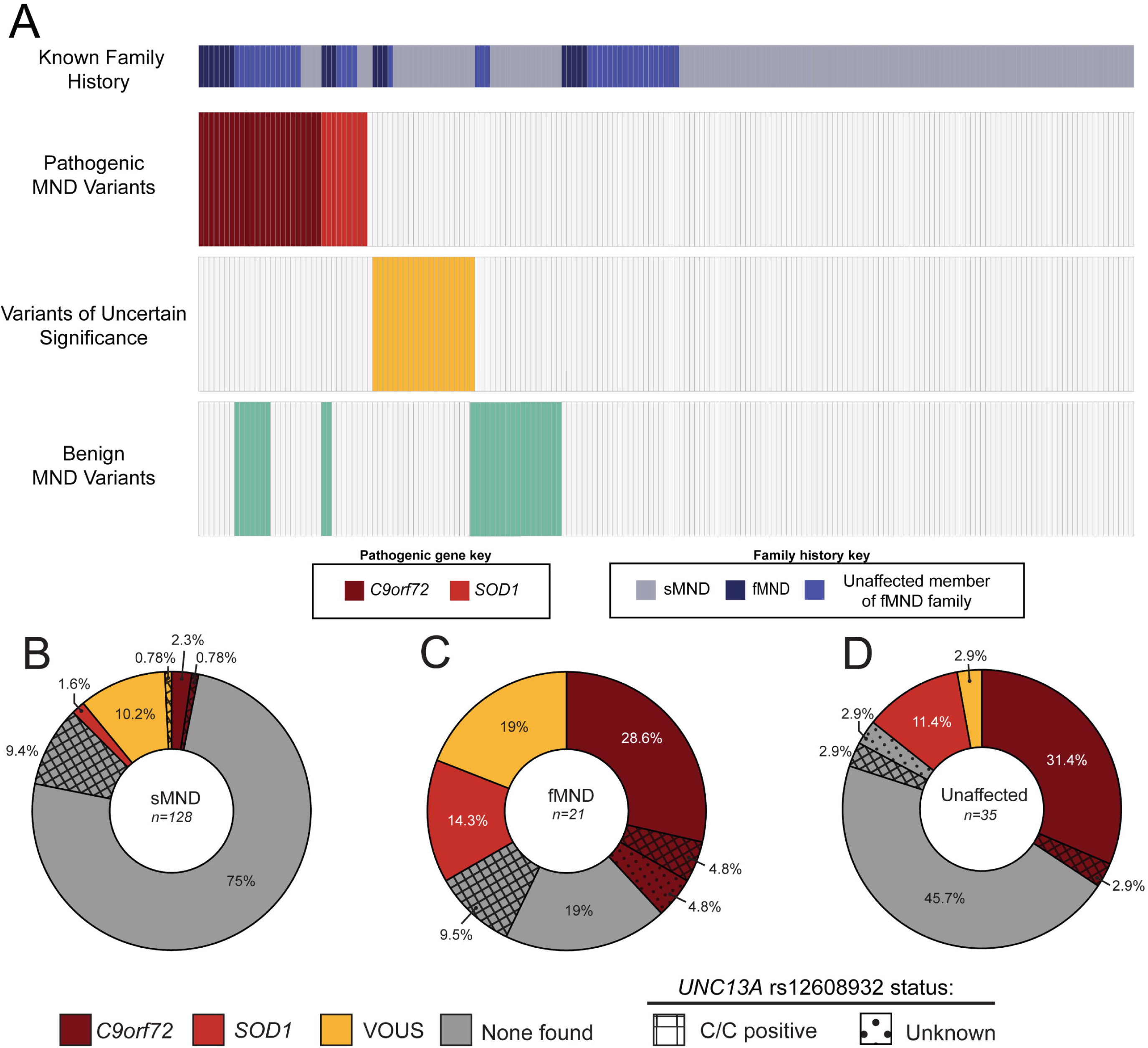
Summary of variants causative of and associated with MND identified in the NZ MND genetics study cohort. A total of 87 MND-associated variants were identified within the cohort, ranging from benign to pathogenic according to ACMG classification. Each column in a given row represents the same participant, showing family history and classes of variant/s carried by that individual (A). These variants accounted for different proportions of participants with sMND (B), fMND (C), and unaffected family members (D). Variants are indicated by colours, with *UNC13A* rs12608932 C/C or (unknown) genotype overlaid. Pathogenic variants were identified in a total of 33 individuals. The most common of these was the *C9orf72* hexanucleotide expansion; (4/128 sporadic cases (3.1%)) (B), 8/21 familial cases (38.2%) (C), and 12/35 unaffected family (35%) (D). Of these, 25% of sporadic *C9orf72* cases, 12.5% of familial *C9orf72* cases and 8.3% of unaffected *C9orf72* cases also harboured the *UNC13A* rs12608932 C/C variant. *SOD1* variants were the next most common pathogenic variants, comprising the p.(Ile114Thr) and p.(Glu101Gly) variants (2/128 sporadic cases (∼1.6%), 3/21 familial cases (14.3%), and 4/35 unaffected family (11.8%)). A total of 14 variants of uncertain significance were also identified in the cohort.

### 1.3.2 *C9orf72* and *SOD1* variants are the most common genetic causes of MND in New Zealand

A total of 87 variants previously reported to be MND-associated (McCann et al., 2020) were identified across 20 genes (Fig. 2A). Pathogenic *C9orf72* NM_001256054.1:c.- 45+163GGGGCC[>60] hexanucleotide repeat expansions were identified in 24 participants: (4 sMND, 8 fMND all from different families, and 12 unaffected family members from 6 different families) (Table 2). Pathogenic autosomal dominant *SOD1* variants were found in nine heterozygous participants: The NM_000454.5:c.341T>C, p.(Ile114Thr) variant was found in five participants (2 sMND, 2 fMND, and 1 related unaffected) and the NM_000454.5:c.301A>G, p.(Glu101Gly) variant was found in four heterozygous participants (1 fMND and 3 related unaffected all within a single family) (Table 2). One uncommon autosomal recessive *SPG7* pathogenic variant was also found in 1 heterozygous participant: NM_003119.4:c.1045G>A, p.(Gly349Ser) (n=1, fMND). Variants that were classified as likely pathogenic were found in 3 heterozygous participants, however, they were either autosomal recessive variants (*SOD1* NM_000454.5:c.272A>C, p.(Asp91Ala) (n=1 sMND)) or likely pathogenic for Paget’s disease of bone rather than MND (*SQSTM1* NM_003900.5:c.1175C>T, p.(Pro392Leu) (n=1 sMND, n=1 fMND)) (Table 2). Likely benign or benign variants that have previously been shown to be associated with MND accounted for 40 of the detected variants (Table 2). Of these, the most common were *OPTN* NM_001008212.2:c.293T>A, p.(Met98Lys) (n=6) and *CFAP410* NM_001271440.1:c.172G>T, p.(Val58Leu) (n=6) (Fig. 2A).

**Table 2.** Variants causative of and associated with MND identified in the NZ MND genetics study cohort.

| Gene | DNA Change | Protein Change | Number of identified samples |  |  |  | Minor allele frequency |  | SNP ID | ClinVar ID | ACMG Class. |
| --- | --- | --- | --- | --- | --- | --- | --- | --- | --- | --- | --- |
|  |  |  | sMND | fMND | Unaffected Family | Total | gnomAD (v.4.1) (NFEU) | ALFA (EUR) |  |  |  |
| *C9orf72* | c.-45+163GGGGGCC[>24] | - | 4 | 8 | 12 | 24 | - | - | - | 31151 | Pathogenic |
| *SOD1* | c.302A>G | p.(Glu101Gly) | 0 | 1 | 3 | 4 | - | - | rs121912439 | 14761 | Pathogenic |
| *SOD1* | c.341T>C | p.(Ile114Thr) | 2 | 2 | 1 | 5 | 2.54E-05 | - | rs121912441 | 197145 | Pathogenic |
| *SPG7* | c.1045G>A | p.(Gly349Ser) | 0 | 1 | 0 | 1 | 2.04E-03 | 1.73E-03 | rs141659620 | 6819 | Pathogenic* |
| *SOD1* | c.272A>C | p.(Asp91Ala) | 1 | 0 | 0 | 1 | 6.10E-04 | 4.53E-04 | rs80265967 | 14766 | Likely Pathogenic * |
| *SQSTM1* | c.1175C>T | p.(Pro392Leu) | 1 | 1 | 0 | 2 | 1.47E-03 | 1.40E-03 | rs104893941 | 8108 | Likely Pathogenic ** |
| *CHCHD10* | c.100C>T | p.(Pro34Ser) | 0 | 1 | 0 | 1 | 2.53E-03 | 2.14E-03 | rs551521196 | 204291 | Likely Benign |
| *DCTN1* | c.3746C>T | p.(Thr1249Ile) | 3 | 0 | 2 | 5 | 5.93E-03 | 4.20E-03 | rs72466496 | 8402 | Likely Benign |
| *SETX* | c.4660T>G | p.(Cys1554Gly) | 1 | 0 | 0 | 1 | 7.09E-03 | 3.28E-03 | rs112089123 | 260505 | Likely Benign |
| *SQSTM1* | c.712A>G | p.(Lys238Glu) | 1 | 0 | 0 | 1 | 3.55E-03 | 3.60E-03 | rs11548633 | 475403 | Benign |
| *CFAP410* | c.172G>T | p.(Val58Leu) | 2 | 0 | 4 | 6 | 1.15E-02 | 1.01E-02 | rs75087725 | 259585 | Benign |
| *CCNF* | c.2140G>A | p.(Val714Met) | 5 | 0 | 0 | 5 | 1.36E-02 | 1.47E-02 | rs61755288 | 774504 | Benign |
| *KIF5A* | c.2957C>T | p.(Pro986Leu) | 1 | 0 | 2 | 3 | 1.37E-02 | 1.19E-02 | rs113247976 | 240087 | Benign |
| *OPTN* | c.293T>A | p.(Met98Lys) | 2 | 0 | 4 | 6 | 2.81E-02 | 3.81E-02 | rs11258194 | 7099 | Benign |

\* Associated with autosomal recessive MND
\*\* Likely pathogenic for Paget's disease of bone, rather than MND

This meant that of the 128 participants with sMND, four (3.1%) carried a pathogenic *C9orf72* hexanucleotide repeat expansion, two (1.6%) carried known pathogenic *SOD1* gene variants (total sMND with pathogenic variants 4.7%), and 14 (16%) carried variants of uncertain significance (VOUS) (Fig. 2B). Of the 21 participants with fMND, eight (38.2%) carried a pathogenic *C9orf72* hexanucleotide repeat expansion, three (14.3%) carried a pathogenic *SOD1* gene variant (total fMND with pathogenic variants 52.4%), and four (19%) carried VOUS (Fig. 2C). Of the 35 unaffected family members, twelve (34.3%) carried a *C9orf72* hexanucleotide repeat expansion, four (11.4%) carried pathogenic *SOD1* gene variants (total unaffected family with pathogenic variants 45.7%), and one (2.9%) carried a VOUS (Fig. 2D). Of the 33 participants who were identified as carrying pathogenic variants, three (9.1%) participants who were positive for *C9orf72* expansion also harboured the *UNC13A* rs12608932 C/C variant associated with disease susceptibility and shorter survival (Diekstra et al., 2012; Van Es et al., 2009) (Fig. 2B-D).

Ten participants with pathogenic MND variants also harboured other MND-associated gene variants in an oligogenic manner (Table 3). These included 8 of the 24 *C9orf72* expansion- positive participants ((33%), from six families). Of these eight, five had two more variants; one had the likely pathogenic *SQSTM1* p.(Pro392Leu) variant (likely pathogenic for Paget’s disease of bone, not MND), and four had *OPTN* p.(Met98Lys) variants. The third variants in this group were two *CFAP410* p.(Val58Leu) variants, one *KIF5A* NM_004984.4:c.2957C>T, p.(Pro986Leu) variant, one *CCNF* NM_001761.3:c.2140G>A, p.(Val714Met) variant and one *DCTN1* NM_004082.5:c.3746C>T, p.(Thr1249Ile) variant. Two of the *SOD1* p.(Glu101Gly)-positive participants, while from the same family (NZ_FALS001), harboured different oligogenic variants: the aforementioned *KIF5A* p.(Pro986Leu) variant in one participant and the *CHCHD10* NM_213720.3:c.100C>T, p.(Pro34Ser) in the other. Three participants with MND with no identified pathogenic MND variants harboured multiple benign variants or VOUS.

**Table 3.** Oligogenic and polygenic cases within the New Zealand MND genetics study cohort.

| Case # | Category | NZ Family ID | Affected / Unaffected | Sex | Variant 1 |  |  | Variant 2 |  |  | Variant 3 |  |  |
| --- | --- | --- | --- | --- | --- | --- | --- | --- | --- | --- | --- | --- | --- |
|  |  |  |  |  | Gene | cDNA Change | Protein Change | Gene | cDNA Change | Protein Change | Gene | cDNA Change | Protein Change |
| NZ_ID_279 | MND-Familial | NZ_FALS044 | Affected | Male | *C9orf72* | c.-45+163GGGGCC[>60] | - | *SQSTM1* | c.1175C>T | p.(Pro392Leu) |  |  |  |
| NZ_ID_100 | Unaffected Family | NZ_FALS004 | Unaffected | Male | *C9orf72* | c.-45+163GGGGCC[>60] | - | *OPTN* | c.293T>A | p.(Met98Lys) | *CFAP410* | c.172G>T | p.(Val58Leu) |
| NZ_ID_117 | Unaffected Family | NZ_FALS004 | Unaffected | Female | *C9orf72* | c.-45+163GGGGCC[>60] | - | *OPTN* | c.293T>A | p.(Met98Lys) | *CFAP410* | c.172G>T | p.Val58Leu |
| NZ_ID_169 | Unaffected Family | NZ_FALS014 | Unaffected | Female | *C9orf72* | c.-45+163GGGGCC[>60] | - | *OPTN* | c.293T>A | p.(Met98Lys) |  |  |  |
| NZ_ID_157 | Unaffected Family | NZ_FALS014 | Unaffected | Female | *C9orf72* | c.-45+163GGGGCC[>60] | - | *OPTN* | c.293T>A | p.(Met98Lys) |  |  |  |
| NZ_ID_144 | Unaffected Family | NZ_FALS003 | Unaffected | Female | *C9orf72* | c.-45+163GGGGCC[>60] | - | *KIF5A* | c.2957C>T | p.(Pro986Leu) |  |  |  |
| NZ_ID_180 | MND-Sporadic | NZ_FALS034 | Affected | Male | *C9orf72* | c.-45+163GGGGCC[>60] | - | *CCNF* | c.2140G>A | p.(Val714Met) |  |  |  |
| NZ_ID_210 | Unaffected Family | NZ_FALS017 | Unaffected | Male | *C9orf72* | c.-45+163GGGGCC[>60] | - | *DCTN1* | c.3746C>T | p.(Thr1249Ile) |  |  |  |
| NZ_ID_136 | Unaffected Family | NZ_FALS001 | Unaffected | Male | *SOD1* | c.302A>G | p.(Glu101Gly) | *KIF5A* | c.2957C>T | p.(Pro986Leu) |  |  |  |
| NZ_ID_76 | MND-Familial | NZ_FALS001 | Affected | Male | *SOD1* | c.302A>G | p.(Glu101Gly) | *CHCHD10* | c.100C>T | p.(Pro34Ser) |  |  |  |
| NZ_ID_60 | MND-Sporadic | - | Affected | Female | *ATP13A2* | c.2412G>A | p.(Lys804=) | *ATP13A2* | c.3193G>A | p.(Val1065Met) |  |  |  |
| NZ_ID_94 | MND-Sporadic | - | Affected | Male | *MAPT* | c.715G>A | p.(Ala239Thr) | *OPTN* | c.293T>A | p.(Met98Lys) | *SPG11* | c.6175C>T | p.(Arg2059Trp) |
| NZ_ID_42 | MND-Familial | NZ_FALS008 | Affected | Female | *GBF1* | c.829G>C | p.(Ala277Pro) | *LRSAM1* | c.375C>A | p.(Asn125Lys) |  |  |  |

**Table 4.**
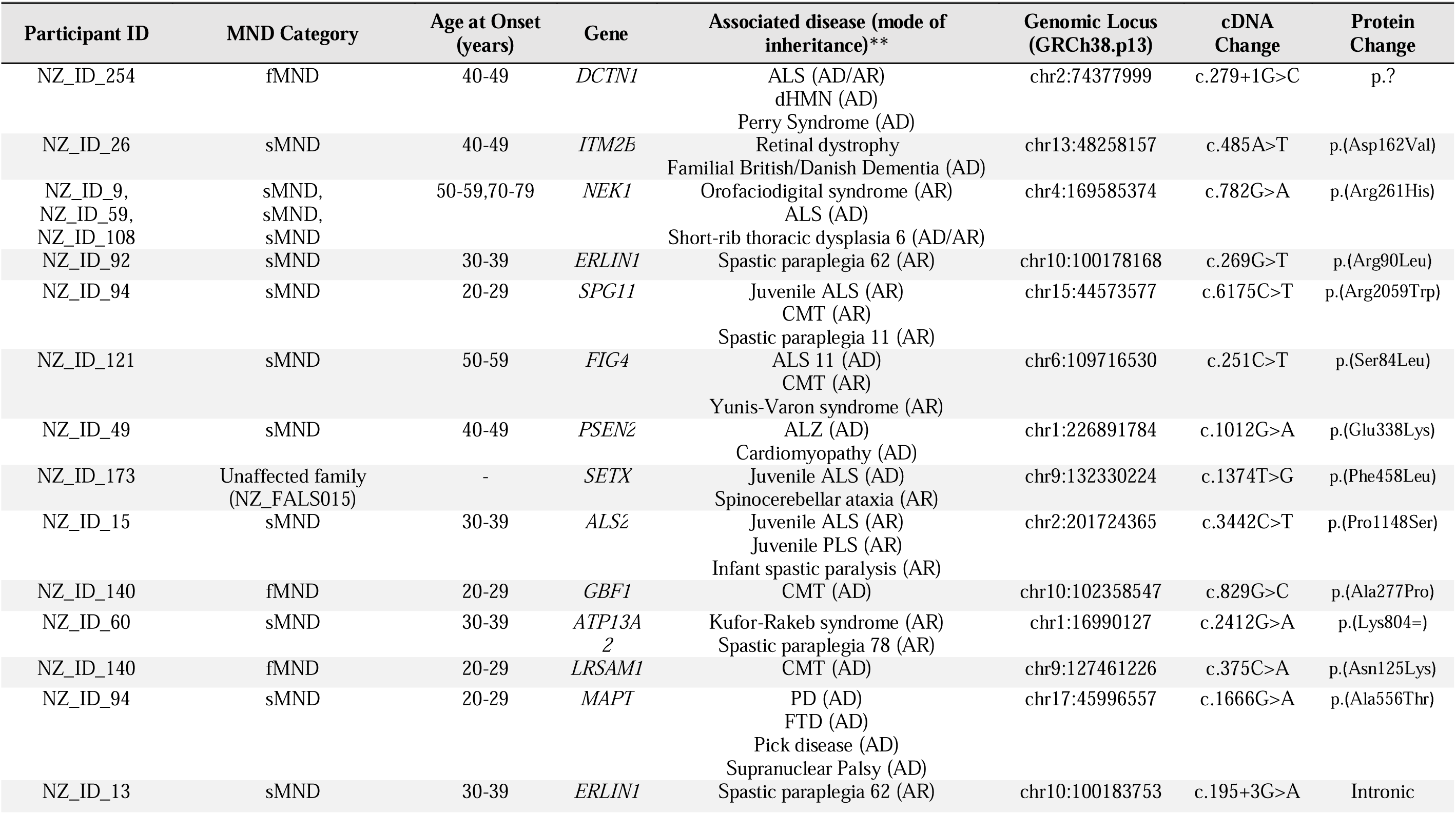
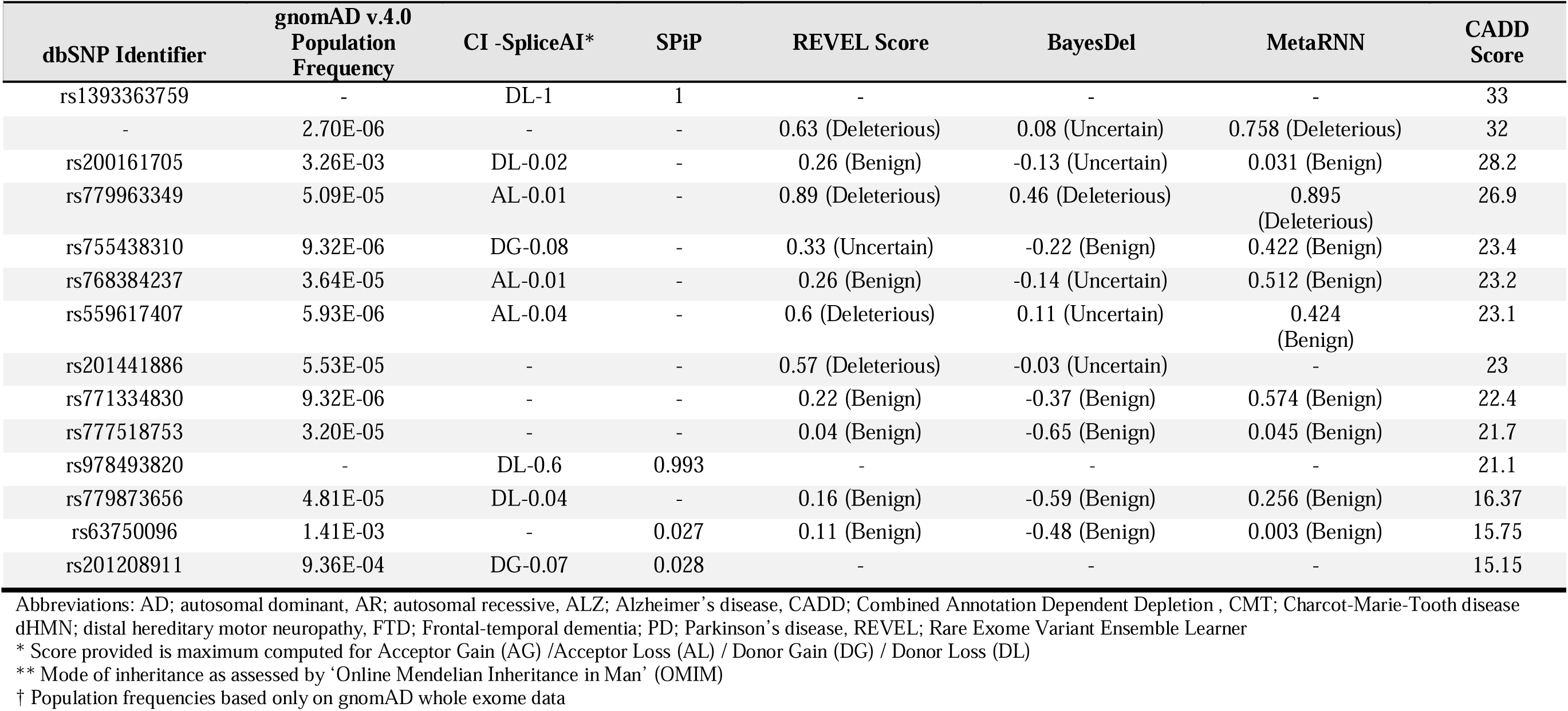
Variants of uncertain significance (VOUS) in the New Zealand MND genetics study cohort.

| Participant ID | MND Category | Age at Onset (years) | Gene | Associated disease (mode of inheritance)** | Genomic Locus (GRCh38.p13) | cDNA Change | Protein Change |
| --- | --- | --- | --- | --- | --- | --- | --- |
| NZ_ID_254 | fMND | 40-49 | *DCTN1* | ALS (AD/AR)<br>dHMN (AD)<br>Perry Syndrome (AD) | chr2:74377999 | c.279+1G>C | p.? |
| NZ_ID_26 | sMND | 40-49 | *ITM2B* | Retinal dystrophy<br>Familial British/Danish Dementia (AD) | chr13:48258157 | c.485A>T | p.(Asp162Val) |
| NZ_ID_9,<br>NZ_ID_59,<br>NZ_ID_108 | sMND,<br>sMND,<br>sMND | 50-59,70-79 | *NEK1* | Orofaciodigital syndrome (AR)<br>ALS (AD)<br>Short-rib thoracic dysplasia 6 (AD/AR) | chr4:169585374 | c.782G>A | p.(Arg261His) |
| NZ_ID_92 | sMND | 30-39 | *ERLIN1* | Spastic paraplegia 62 (AR) | chr10:100178168 | c.269G>T | p.(Arg90Leu) |
| NZ_ID_94 | sMND | 20-29 | *SPG11* | Juvenile ALS (AR)<br>CMT (AR)<br>Spastic paraplegia 11 (AR) | chr15:44573577 | c.6175C>T | p.(Arg2059Trp) |
| NZ_ID_121 | sMND | 50-59 | *FIG4* | ALS 11 (AD)<br>CMT (AR)<br>Yunis-Varon syndrome (AR) | chr6:109716530 | c.251C>T | p.(Ser84Leu) |
| NZ_ID_49 | sMND | 40-49 | *PSEN2* | ALZ (AD)<br>Cardiomyopathy (AD) | chr1:226891784 | c.1012G>A | p.(Glu338Lys) |
| NZ_ID_173 | Unaffected family (NZ_FALS015) | - | *SETX* | Juvenile ALS (AD)<br>Spinocerebellar ataxia (AR) | chr9:132330224 | c.1374T>G | p.(Phe458Leu) |
| NZ_ID_15 | sMND | 30-39 | *ALS2* | Juvenile ALS (AR)<br>Juvenile PLS (AR)<br>Infant spastic paralysis (AR) | chr2:201724365 | c.3442C>T | p.(Pro1148Ser) |
| NZ_ID_140 | fMND | 20-29 | *GBF1* | CMT (AD) | chr10:102358547 | c.829G>C | p.(Ala277Pro) |
| NZ_ID_60 | sMND | 30-39 | *ATP13A2* | Kufor-Rakeb syndrome (AR)<br>Spastic paraplegia 78 (AR) | chr1:16990127 | c.2412G>A | p.(Lys804=) |
| NZ_ID_140 | fMND | 20-29 | *LRSAM1* | CMT (AD) | chr9:127461226 | c.375C>A | p.(Asn125Lys) |
| NZ_ID_94 | sMND | 20-29 | *MAPT* | PD (AD)<br>FTD (AD)<br>Pick disease (AD)<br>Supranuclear Palsy (AD) | chr17:45996557 | c.1666G>A | p.(Ala556Thr) |
| NZ_ID_13 | sMND | 30-39 | *ERLIN1* | Spastic paraplegia 62 (AR) | chr10:100183753 | c.195+3G>A | Intronic |

| dbSNP Identifier | gnomAD v.4.0<br>Population<br>Frequency | CI -SpliceAI* | SPiP | REVEL Score | BayesDel | MetaRNN | CADD<br>Score |
| --- | --- | --- | --- | --- | --- | --- | --- |
| rs1393363759 | - | DL-1 | 1 | - | - | - | 33 |
| - | 2.70E-06 | - | - | 0.63 (Deleterious) | 0.08 (Uncertain) | 0.758 (Deleterious) | 32 |
| rs200161705 | 3.26E-03 | DL-0.02 | - | 0.26 (Benign) | -0.13 (Uncertain) | 0.031 (Benign) | 28.2 |
| rs779963349 | 5.09E-05 | AL-0.01 | - | 0.89 (Deleterious) | 0.46 (Deleterious) | 0.895<br>(Deleterious) | 26.9 |
| rs755438310 | 9.32E-06 | DG-0.08 | - | 0.33 (Uncertain) | -0.22 (Benign) | 0.422 (Benign) | 23.4 |
| rs768384237 | 3.64E-05 | AL-0.01 | - | 0.26 (Benign) | -0.14 (Uncertain) | 0.512 (Benign) | 23.2 |
| rs559617407 | 5.93E-06 | AL-0.04 | - | 0.6 (Deleterious) | 0.11 (Uncertain) | 0.424<br>(Benign) | 23.1 |
| rs201441886 | 5.53E-05 | - | - | 0.57 (Deleterious) | -0.03 (Uncertain) | - | 23 |
| rs771334830 | 9.32E-06 | - | - | 0.22 (Benign) | -0.37 (Benign) | 0.574 (Benign) | 22.4 |
| rs777518753 | 3.20E-05 | - | - | 0.04 (Benign) | -0.65 (Benign) | 0.045 (Benign) | 21.7 |
| rs978493820 | - | DL-0.6 | 0.993 | - | - | - | 21.1 |
| rs779873656 | 4.81E-05 | DL-0.04 | - | 0.16 (Benign) | -0.59 (Benign) | 0.256 (Benign) | 16.37 |
| rs63750096 | 1.41E-03 | - | 0.027 | 0.11 (Benign) | -0.48 (Benign) | 0.003 (Benign) | 15.75 |
| rs201208911 | 9.36E-04 | DG-0.07 | 0.028 | - | - | - | 15.15 |

Abbreviations: AD; autosomal dominant, AR; autosomal recessive, ALZ; Alzheimer's disease, CADD; Combined Annotation Dependent Depletion, CMT; Charcot-Marie-Tooth disease dHMN; distal hereditary motor neuropathy, FTD; Frontal-temporal dementia; PD; Parkinson's disease, REVEL; Rare Exome Variant Ensemble Learner
\* Score provided is maximum computed for Acceptor Gain (AG) / Acceptor Loss (AL) / Donor Gain (DG) / Donor Loss (DL)
\*\* Mode of inheritance as assessed by 'Online Mendelian Inheritance in Man' (OMIM)
† Population frequencies based only on gnomAD whole exome data

### 1.3.3 A novel *DCTN1* variant and 13 other variants of uncertain significance may be contributors to genetic risk of MND in New Zealand

Variants of uncertain significance were parsed through *in silico* pathogenicity predictive tools (Table 3). A total of 14 different variants, found in 14 participants, remained following CADD-based filtering, with the highest scores seen for *DCTN1* NM_004082.5:c.279+1G>C p.(?) (CADD: 33), *ITM2B* NM_021999.5:c.485A>T, p.(Asp162Val) (CADD: 32, MetaRNN: 0.76, REVEL: 0.63), *NEK1* NM_001199397.3:c.782G>A, p.(Arg261His) (CADD: 28.2, MetaRNN: 0.031, REVEL: 0.26), and *ERLIN1* NM_006459.4:c.269G>T, p.(Arg90Leu) (CADD: 26.9, MetaRNN: 0.895, REVEL: 0.89), but their pathogenicity has yet to be experimentally validated.

Both the *DCTN1* NM_004082.5:c.279+1G>C p.(?) and *ATP13A2* NM_022089.4:c.2412G>A, p.(Lys804=) variants were identified by both CI-SpliceAI (donor loss scores of 1 and 0.6 respectively ) and SPiP (1 and 0.993, respectively) algorithms as being likely to impact normal splicing, resulting in the loss of a splice donor site. However, these predictions need to be tested empirically.

### 1.3.4 Identity-by-descent analysis identifies four New Zealand participants sharing a *SOD1* founder haplotype with >30 Australian ALS cases

Two apparently sporadic New Zealand ALS participants who carried the *SOD1* NM_000454.5:c.341T>C, p.(Ile114Thr) variant were identified as being identical-by-descent over the *SOD1* gene locus to Australian familial and sporadic cases, who in turn have been shown to be related to a larger cluster of >30 MND cases carrying the same variant (Fig. 3). All of these participants share a founder disease haplotype (Henden et al., 2020). All related New Zealand-Australian pairs were estimated to be >7^th^ degree relatives. Additionally, two other cases (n=1, fMND; n=1, unaffected family) were linked to this wider Australian *SOD1* cluster.

**Figure 3.**
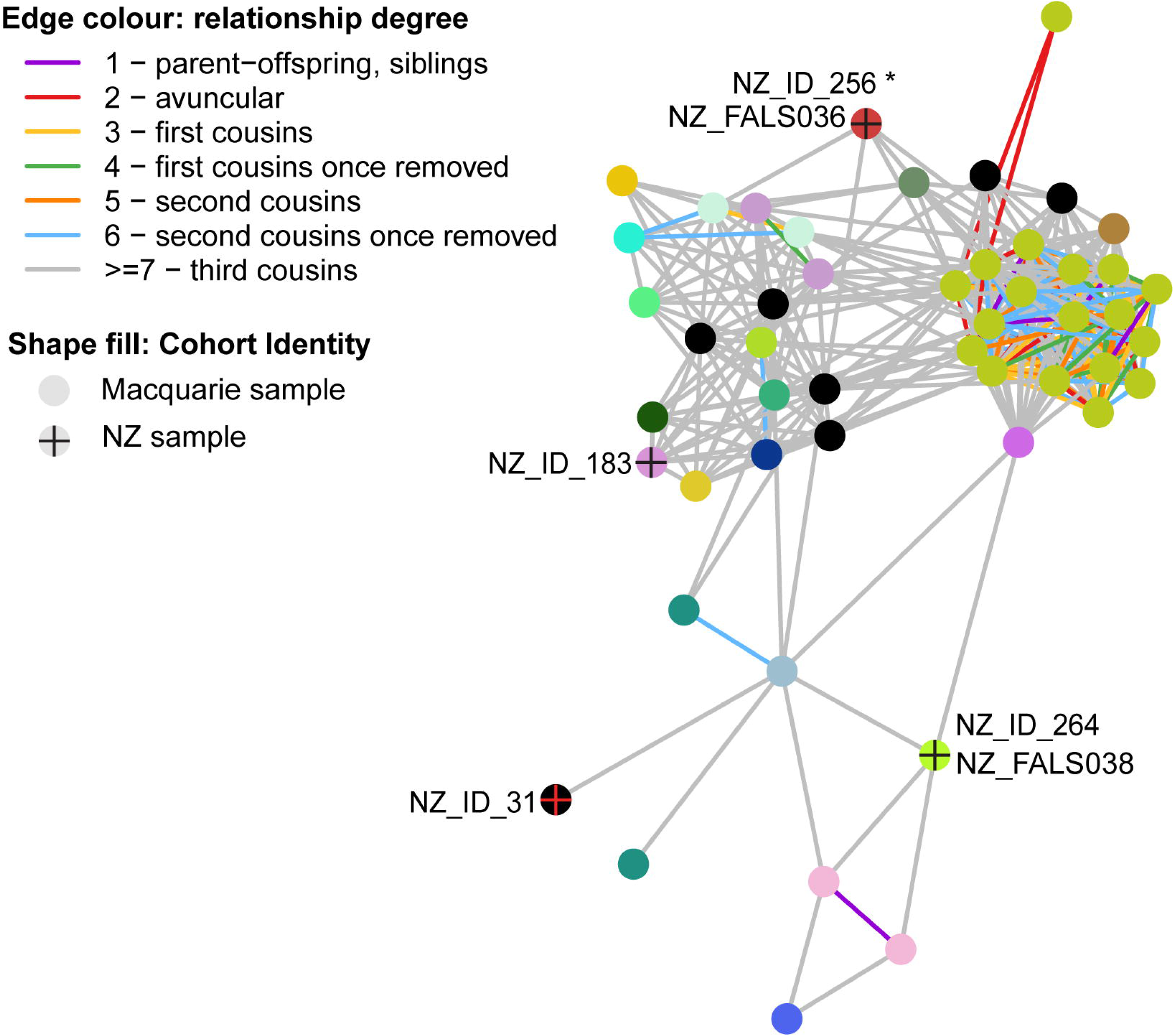
Identification of four New Zealand *SOD1* p.(Ile114Thr) cases linked to a wider Australian *SOD1* p.(Ile114Thr) network using identity-by-descent analysis. Four New Zealand cases harbouring a *SOD1* p.(Ile114Thr) variant (2 sMND, 1fMND, and 1 unaffected family) were identified as ‘identical-by-descent’ to a cluster of >30 and familial and sporadic MND cases from Australia with the same variant. All relationships involving New Zealanders were estimated to be >7th degree relative pairings. Links (lines) represent identity-by- descent across a locus of interest, in this case *SOD1,* with link colour denoting degree of relatedness. Asterix (*) denotes unaffected individual

## 1.4 Discussion

The development of therapies targeting MND is increasingly focussed on gene-specific approaches, necessitating genetic testing as a tool to assess patient eligibility. New Zealand, despite having among the highest mortality rates of MND in the world (Cao et al., 2018; Logroscino et al., 2018), has conducted little genetic research of its MND population. This study aimed to address the paucity of information surrounding the genetic determinants of MND in New Zealand through genetic testing of clinically affected and at-risk participants and thus empower individuals to seek clinical trial enrolment and/or treatment with approved gene-specific drugs.

Our New Zealand MND cohort consisted of 184 participants with clinical phenotypes that were comparable to a European MND reference population. While we did not see any differences in the age of disease onset within our cohort between sex, familial status, and site of onset, this is likely due to low sample size rather than a reflection of differences from the reference cohort in underlying biology.

Causative autosomal dominant variants were identified in 33 participants (24 *C9orf72*, 9 *SOD1)*. Twelve of these individuals were unaffected family members. Identity-by-descent analysis was used to examine the penetrance of a pathogenic SOD1 p.(Ile114Thr) variant, finding that two participants with apparently sporadic MND carrying this variant shared the variant haplotype with 31 familial and sporadic cases MND cases in Australia. This variant is among the most frequent MND-causing *SOD1* variants in UK populations but shows variable penetrance, supported here by our participants reporting no known family history of MND (Aggarwal and Nicholson, 2005; Cudkowicz et al., 1997; Hayward et al., 1996; Lopate et al., 2010; McCann et al., 2017; Shepheard et al., 2021; Spargo et al., 2022).

In addition to those with autosomal dominant pathogenic variants, there were three participants who harboured pathogenic or likely pathogenic variants that were unlikely to have caused their MND. Each of these variants (*SQSTM1* p.(Pro392Leu), *SPG7* p.(Gly349Ser), and *SOD1* p.(Asp91Ala)) was carried heterozygously. The *SQSTM1* p.(Pro392Leu) variant is frequent in European populations (Non-Finnish European (NFEU): MAF: 1.42×10^-3^, gnomAD v4.0.0) and is pathogenic in the heterozygous state, not for MND but for Paget’s disease of bone (*OMIM*: 167250). Individuals with MND harbouring this variant typically present with concomitant Paget’s disease, suggesting disease segregation with Paget’s disease rather than with MND (Almeida et al., 2016; Le Ber, 2013; Teyssou et al., 2013).

*SPG7* is associated with autosomal recessive hereditary spastic paraplegia 7 which is characterised by progressive bilateral lower limb spasticity (Harding, 1983). The *SPG7*, p.(Gly349Ser) variant, detected in one participant in this study, was also detected in a large Dutch cohort in 7 of 728 participants (0.96%), all of whom were compound heterozygotes, and in 0.3% of controls, consistent with the variant being classified as autosomal recessive (Van Gassen et al., 2012). The variant is also present in European reference populations (NFEU MAF: 2.04×10^-3^, gnomAD v4.0.0) with few homozygotes. No other *SPG7* variants were detected in our participant, so whether this variant conferred susceptibility to their disease is unclear.

The *SOD1* p.(Asp91Ala) variant is the most common *SOD1* variant identified in European populations but reports on its pathogenicity are conflicting (Forsberg et al., 2023). The variant is typically classified as autosomal recessive, particularly in northern Scandinavian countries like Finland where it is particularly prevalent (NFEU MAF: 6.13×10^-4^ vs. Finnish European (FEU) MAF: 0.017, gnomAD v4.0.0) (Jonsson et al., 2002; M Andersen, Victor A Spitsyn, Sergu, 2001; Parton et al., 2002). However, in other European countries it has been reported as autosomal dominant; being reported in 3 British sporadic cases (Jackson et al., 1997) and in 2 Belgian familial ALS cases (Robberecht et al., 1996), all five heterozygous and the two familial cases showing segregation with disease.

Two VOUS identified in our study (*DCTN1:*c.279+1G>C p.(?) and *ATP13A2* p.(Lys804=)) are subject to ongoing study for potential effects on splicing, given that they are both predicted to alter donor splice sites. We identified 11 other VOUS in unrelated participants, some of which warrant further investigation by virtue of their *in silico* predicted deleteriousness such as *ERLIN1* p.(Arg90Leu) and *ITM2B* p.(Asp162Val). ER Lipid Raft Associated 1 (*ERLIN1)* variants have mainly been implicated in hereditary spastic paraplegia (Novarino et al., 2014; Zhu et al., 2022) but a homozygous NM_006459.4:c.281T>C, p.(Val94Ala) variant was described in early-onset MND (Tunca et al., 2018). Similarly, both dominant and recessive variants in the paralog *ERLIN2* have been associated with initial spastic paraplegia rapidly progressing to MND (Amador et al., 2019). Integral membrane protein 2B (*ITM2B/BRI2)* is typically associated with familial British and Danish dementia wherein autosomal dominant variants result in the production of longer amyloidogenic peptides (Ghiso et al., 2006; Yao et al., 2019). In MND, *ITM2B* variants have been found in several Italian cases; one ALS/ FTD case carrying a NM_021999.5:c.751A>G, p.(Ile251Val) variant together with a *C9orf72* expansion, and two sALS cases with a c.511A>C, p.(Thr171Pro) variant (Bartoletti-Stella et al., 2021; Giannoccaro et al., 2018). Interestingly, the p.(Thr171Pro) variant lies within the functionally important BRICHOS domain (residues 137-231) of ITM2B (Leppert et al., 2023), as does the p.(Asp162Val) variant identified in our study which may suggest a common mechanism of pathogenicity. The risk variant *UNC13A* NM_001080421.3:c.2473-324A>C p.(?), was present as homozygous in 8.3% and is associated with shorter survival compared to A/A or A/C genotypes (Diekstra et al., 2012; Gaastra et al., 2016; Tan et al., 2020; Van Es et al., 2009; Vidal-Taboada et al., 2015; Willemse et al., 2023).

Three benign variants, *OPTN* p.(Met98Lys), *CFAP410* p.(Val58Leu), and *KIF5A* p.(Pro986Leu), identified in 6 cases in our cohort, together with pathogenic variants in another gene (*C9orf72* n=5, *SOD1* n=1) have also been reported in other MND cohorts (Brenner et al., 2018; Bury et al., 2016; McCann et al., 2020; PARALS Registry et al., 2016). Their presence may reduce the liability threshold required for MND manifestation and thus contribute to a pattern of oligogenic inheritance (Brenner et al., 2018; Bury et al., 2016; McCann et al., 2020; van Blitterswijk et al., 2012).

The overall contribution of MND genes to risk of MND in New Zealand could not be ascertained because enrolment in our study was through self-selection (non-random sampling of New Zealanders with sporadic and familial MND) and included at-risk unaffected family members of people with fMND. However, the overall finding of pathogenic variants in 4.7% of sMND, 52.4% of fMND, and 45.7% of unaffected family members, most commonly in *C9orf72* and *SOD1,* will inform New Zealand’s approach to genetic testing and research in MND. Currently in New Zealand, free genetic testing for people with, or at risk for, MND is offered through the public health service at their neurologist’s request or via a referral to the genetic health service, respectively. However, those with MND depend upon individual neurologists requesting appropriate tests, and those at risk face long wait times and strict acceptance criteria for these genetic tests. In practice, most New Zealanders with sMND are not offered genetic testing. Similarly in the US, Canada, UK, and Europe, current guidelines for genetic testing in MND primarily reflect the outdated distinction between fMND and apparent sMND (Miller et al., 2009; National Institute for Health and Care Excellence, 2016;

Salmon et al., 2022; Shoesmith et al., 2020; The EFNS Task Force on Diagnosis and Management of Amyotrophic Lateral Sclerosis: et al., 2012). There is increasing recognition amongst researchers and clinicians that genetic testing should be available to everyone with MND, regardless of apparent inheritance (Dharmadasa et al., 2022; Dilliott et al., 2023; Mehta et al., 2022; Roggenbuck et al., 2023; Salmon et al., 2022; Shepheard et al., 2021) and which this research supports.

Accessible genetic testing is of particular importance now that gene-specific treatments are available and continue to be developed for MND. The antisense oligonucleotide Tofersen (Qalsody, Biogen) was recently conditionally approved by the FDA for ALS in adults with *SOD1*-linked ALS and is subject to an ongoing trial for pre-symptomatic administration (ATLAS; NCT04856982) (Benatar et al., 2022). Similarly, post-hoc meta-analyses suggest that survival in people with MND carrying the *UNC13A* rs12608932 C/C variant can be extended by treatment with lithium carbonate (van Eijk et al., 2017), which is safe and inexpensive and the subject of an ongoing clinical trial (MAGNET; EudraCT- 2020_000579_19) (Willemse et al., 2023). Eligibility for treatment with gene-specific therapies or entry into their clinical trials requires prior knowledge of gene variant status, thus highlighting the need to consider early genetic testing in all New Zealanders diagnosed with MND and their at-risk family members, to enable early therapeutic intervention. Within our study, we uncovered 21 individuals who could be eligible for *SOD1* or *UNC13A* clinical trials by virtue of knowing their genetic test results.

Overall, our study has investigated the genetic heterogeneity of MND in New Zealand. Our data suggest that the genetics of MND in our New Zealand cohort largely resembles that in European populations, being most commonly caused by pathogenic genetic variants in *C9orf72* or *SOD1*. However, New Zealand is also home to additional ethnic populations with MND (including, Asian, NZ Māori, and Pasifika) with whom we seek to engage further to explore genetic causation. Given the number of pre-disposed carriers identified, and the finding of gene-positive ‘sporadic’ cases including two related to a large MND cluster, our study supports the notion that genetic screening services should be free for all New Zealanders with MND, and that at-risk family members could also benefit from gene-specific therapies.

## 1.5 Author Contributions

ELS and RR conceptualised the study. ELS, JD, SK, SN, and CV liaised with and recruited participants for the study. MR, CB, and HF gathered informed participant consent and returned results to participants. DM collated information from the MND registry. HW, KD, EP, and HP processed participant blood and saliva, and conducted DNA extraction and accredited Sanger and *C9orf72* fragment analysis/ RP-PCR testing. AH oversaw research-grade Illumina GSA, Sanger sequencing, *C9orf72* fragment analysis and RP-PCR testing. EM helped conduct GSA processing. LH and KW conducted the identity-by-descent analysis. Data processing, analysis, and interpretation were carried out by MM, JJ, RR, and ELS. Manuscript was prepared by MM and ELS and reviewed by all authors.

## 1.6 Declarations and Interests

### 1.6.3 Ethics approval

All participants in the study gave their informed consent. The study was approved by the Health and Disability Ethics Committee (HDEC approval number 19/CEN/7).

## Data Availability

Non-identifiable data produced in the present study are available upon reasonable request to the authors.

## Acknowledgements

This publication is dedicated to the incredible participants and families who contribute to our research. The authors would also like to thank the ‘Project MinE GWAS Consortium’ for the use of their clinical data.

