## Supplementary for "Genetics of Motor Neuron Disease in New Zealand"

**SUPPLEMENTARY MATERIALS**

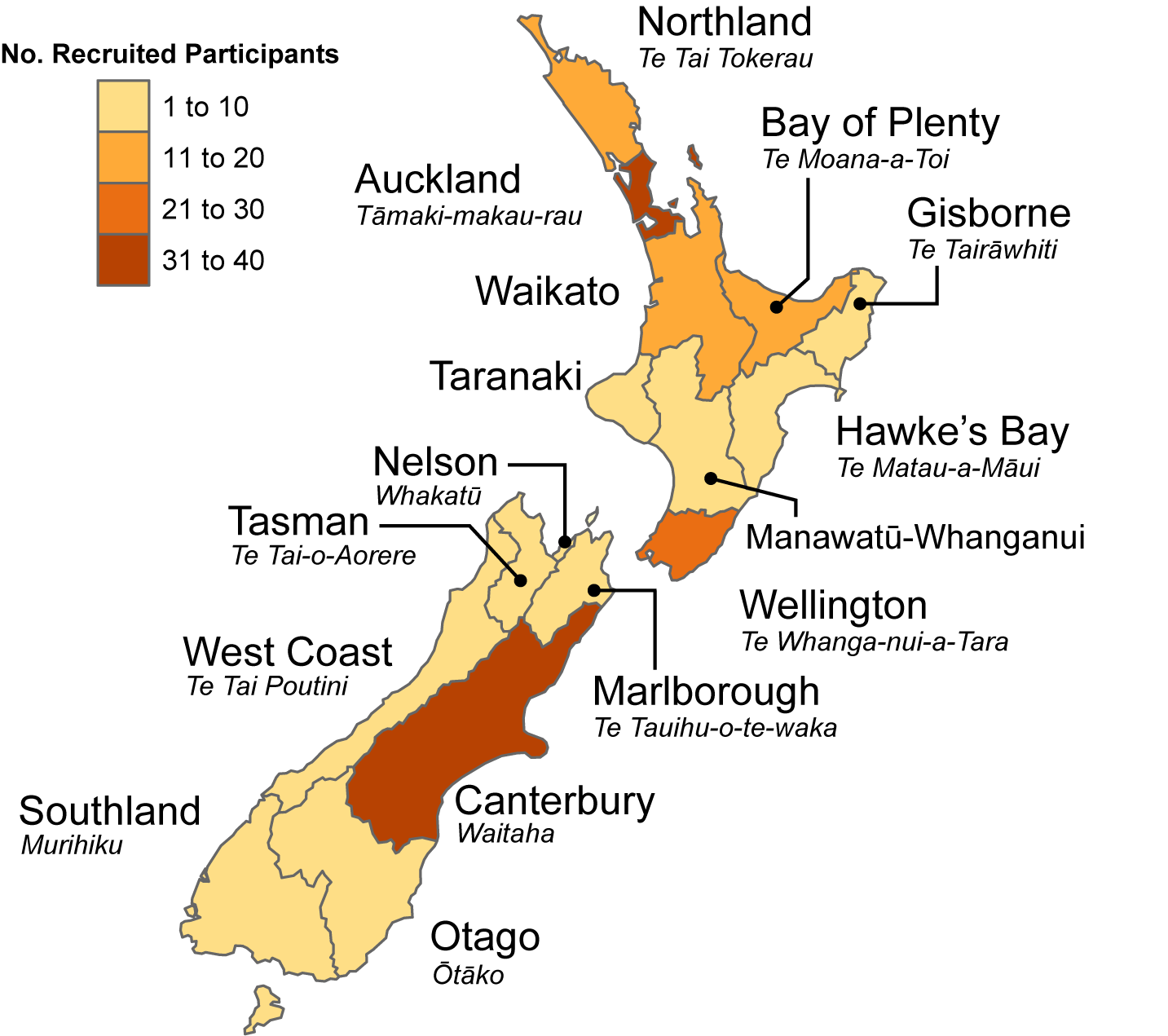

**Supplementary Fig. 1: Geographical distribution of participants recruited in the New Zealand MND cohort.** Participants were recruited from all 16 regions of New Zealand with Auckland, Canterbury, and Wellington being the most represented regions with 38, 34, and 26 participants respectively, which aligns with these being major population centres. Geographic data was available for 179/185 participants.

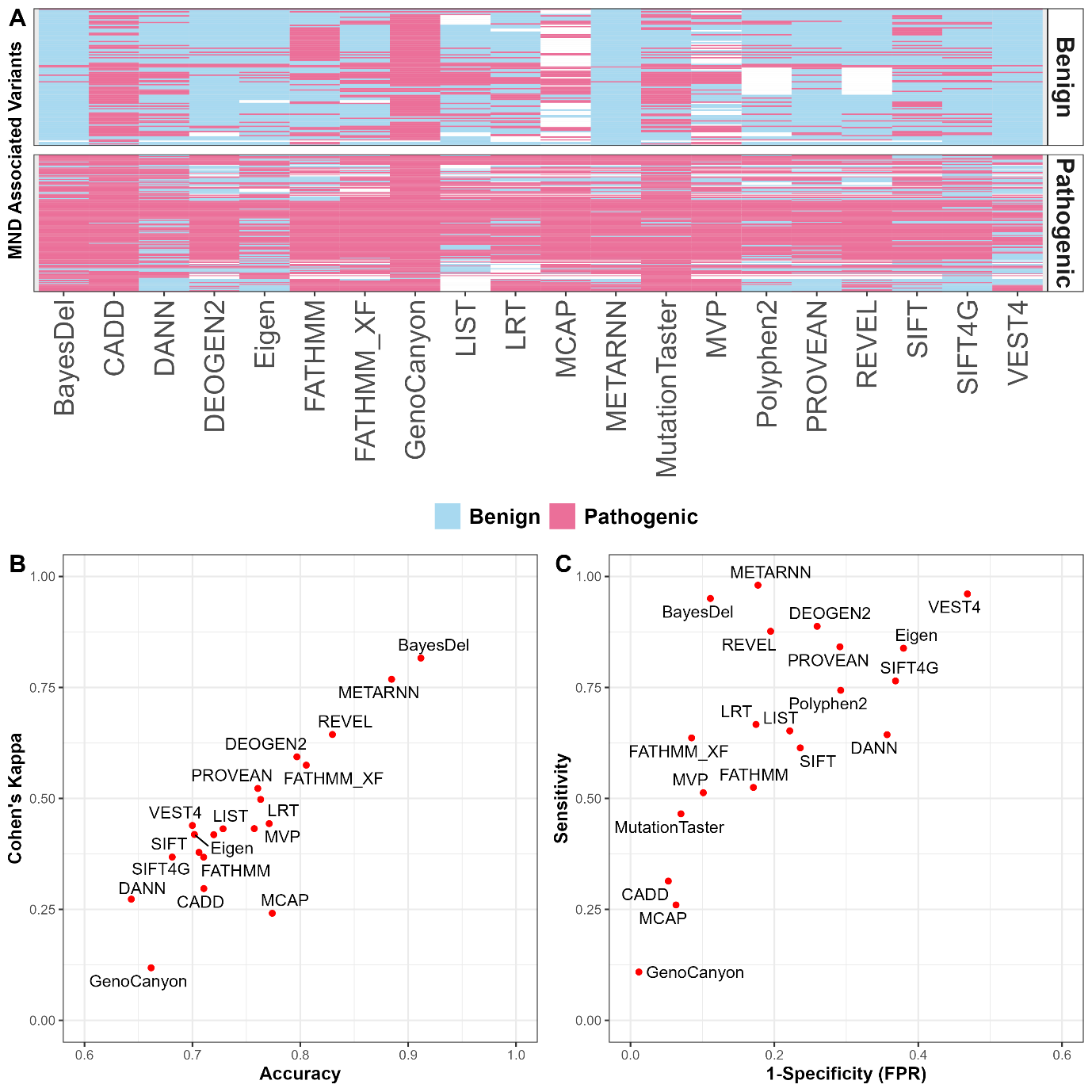

**Supplementary Fig. 2: Missense variant predictor accuracy evaluation using ClinVar MND associated variants.**

**A)** Visualisation of algorithm predictions with ‘ground-truth’ MND associated variants (n=237) in the two vertical boxes and with predictions coloured as either blue (benign) or pink (pathogenic). Cohen’s kappa for inter-rater reliability between prediction and ground truth was computed and compared with accuracy **(B)** and false positive rate **(C).** In both instances, BayesDel and MetaRNN seemed to perform the best (Cohen’s Kappa > 0.75) while GenoCanyon consistently over-predicted deleteriousness.

| **Supplementary Table 1:** Variants analysed by Sanger sequencing | | | |
| --- | --- | --- | --- |
| **Gene** | **dbSNP ID** | **Mutation Location (hg19)** | **Protein mutation** |
| *FUS* | rs121909668 | 16:31202739C>T,A | p.R521C,S |
|  | rs121909671 | 16:31202740G>A | p.R521H |
|  | rs267606831 | 16:31202410G>A | p.G507D |
| *SOD1* | rs80265967 | 21:33039603A>C,T | p.D91A,V |
|  | rs121912438 | 21:33039612G>T,C,A | p.G94V,A,D |
|  | rs121912441 | 21:33039672T>C | p.I114T |
| *TARDBP* | rs80356719 | 1:11082325G>A | p.G287S |
|  | rs753730998 | 1:11082593G>A | p.G376D |
|  | - | 1:11082623delAT insCACCAACC | p.S387delinsTNP |
|  | rs80356721 | 1:11082347G>C | p.G294A |
|  | rs80356727 | 1:11082457C>A | p.Q331K |
|  | rs80356730 | 1:11082475A>G | p.M337V |
| *C9orf72* | rs3849942 | 9:27543281T>A | - |
| *UNC13A* | rs12608932 | 19:17752689A>C,T | - |

| **Supplementary Table 3:** Invitae combined ALS/FTD gene panel | |
| --- | --- |
| **Gene** | **Transcript** |
| *ALS2* | NM_020919.3 |
| *ANG* | NM_001145.4 |
| *ANXA11* | NM_001157.2 |
| *APP* | NM_000484.3 |
| *ATP13A2* | NM_022089.3 |
| *CHCHD10* | NM_213720.2 |
| *CHMP2B* | NM_014043.3 |
| *DCTN1* | NM_004082.4 |
| *DDHD1* | NM_001160147.1 |
| *ERBB4* | NM_005235.2 |
| *ERLIN1* | NM_006459.3 |
| *FIG4* | NM_014845.5 |
| *FUS* | NM_004960.3 |
| *GRN* | NM_002087.3 |
| *HEXA* | NM_000520.4 |
| *HNRNPA2B1* | NM_031243.2 |
| *ITM2B* | NM_021999.4 |
| *KIF5A* | NM_004984.2 |
| *LRRK2* | NM_198578.3 |
| *MAPT* | NM_005910.5 |
| *MATR3* | NM_199189.2 |
| *NEFH* | NM_021076.3 |
| *OPTN* | NM_021980.4 |
| *PFN1* | NM_005022.3 |
| *PRNP* | NM_000311.3 |
| *PSEN1* | NM_000021.3 |
| *PSEN2* | NM_000447.2 |
| *SETX* | NM_015046.5 |
| *SIGMAR1* | NM_005866.3 |
| *SNCA* | NM_000345.3 |
| *SOD1* | NM_000454.4 |
| *SORL1* | NM_003105.5 |
| *SPG11* | NM_025137.3 |
| *SQSTM1* | NM_003900.4 |
| *TARDBP* | NM_007375.3 |
| *TBK1* | NM_013254.3 |
| *TFG* | NM_006070.5 |
| *TIA1* | NM_022173.2 |
| *TREM2* | NM_018965.3;NM_001271821.1 |
| *UBQLN2* | NM_013444.3 |
| *VAPB* | NM_004738.4 |
| *VCP* | NM_007126.3 |

| **Supplementary Table 4:** Primers used for *C9orf72* RP-PCR and fragment analysis | | |
| --- | --- | --- |
|  | **Primer Name** | **Sequence** |
| **RP-PCR** | C9orf72_F_FAM (Primer 1 | FAM-AGTCGCTAGAGGCGAAAGC |
|  | C9orf72_R (Primer 2) | tacgcatcccagtttgagacgGGGGCCGGGGCCGGGGCCGGGG |
|  | C9orf72_Anchor (Primer 3) | tacgcatcccagtttgagacg |
| **Fragment Analysis** | C9orf72_FIRST_R_FAM (Primer 4) | FAM-CAAGGAGGGAAACAACCGCAGCC |
|  | C9orf72_FIRST_F (Primer 5) | GCAGGCACCGCAACCGCAG |

| **Supplementary Table 5:** Pathogenicity cut-off scores for various *in silico* pathogenicity prediction algorithms | |
| --- | --- |
| **Algorithm score used:** | **Pathogenicity cutoff score** |
| SIFT_pred | >0.39575 |
| SIFT4G_score | < 0.05 |
| Polyphen2_HVAR_rankscore | >0.447 |
| LRT_pred | see dbNSFP |
| MutationTaster_pred | >0.31733 |
| FATHMM_pred | ≥0.81332 |
| PROVEAN_score | ≥0.54382 |
| VEST4_score | ≥0.8 * |
| MetaRNN_score | >0.6149 |
| M_CAP_score | >0.025 |
| REVEL_score | >0.5 |
| MVP_score | >0.75 |
| DEOGEN2_score | >0.5 |
| BayesDel_addAF_score | >0.0692655 |
| LIST_S2_score | >0.85 |
| CADD_phred | >15 |
| DANN_score | ≥0.993 |
| fathmm_XF_coding_score | >0.5 |
| Eigen_raw_coding | >0.5 |
| GenoCanyon_score | >= 0.5 |

* Inferred from (Carter et al., 2013; Quinodoz et al., 2022)
